# Can we see gender through the microscope? The research output using menstrual blood cells has much to say

**DOI:** 10.1101/2022.02.15.22270947

**Authors:** Daniela Tonelli Manica, Karina Dutra Asensi, Gaia Mazzarelli, Germana Barata, Regina Coeli dos Santos Goldenberg

## Abstract

Despite proven scientific quality of menstrual blood mesenchymal cells, research and science output using those cells is still incipient and considered taboo. This study analyzes the literature of the menstrual blood mesenchymal stromal/stem cells (mbMSC) at PubMed database between (2008 – 2020) and the social attention it received on Twitter. A comparative analysis showed that mbMSC has a very small space within mesenchymal cells research (0.25%). Most first authors are women (53.2%), whereas most last authors are men (63.74%). Menstrual blood tends to be less used in experiments and its scientific value tends to be underestimated, which brings gender bias to a technical and molecular level. Although women are more positive in the mbMSC debate on Twitter, communication efforts toward visibility and public interest in menstrual cells has room to grow.

## Introduction

Studies in the last half century have broadly considered gender issues in science (1, 2, 3, 4, 5, 6, 7, 8). Feminist and postcolonial approaches to science have recently shown the complex interrelations between inequalities of gender, race/ethnicity, and class within scientific practices (9, 10). The presence of gender and racial bias in science is well-known, and many studies focus on the presence and proportion of women and other minorities in different social contexts within laboratories and universities.

Recent efforts have been trying to increase the visibility of the usual social dynamics that limit women and other minorities’ scientific career, sometimes articulated to social movement’s political agendas (#metoo, #blacklivesmatter). Reports show that women tend to have lower remuneration and peer acknowledgment, and that there is a bottleneck, or a glass ceiling, for women and other minorities to achieve a higher status in academic and other power hierarchies (11). Demands over women’s evaluation tend to be higher (12), and experiments with non-blind reviews have shown that they tend to favor white men (13). Male authors may be associated with greater scientific quality (14) and gender bias influences the review process (15). Furthermore, gender and international diversity are related to lower acceptance rates in peer reviews (16).

In November 2020, the publication of a paper by Nature Communications has shaken the scientific community (17). Based on an analysis of shared authorship in papers and citation rates, the authors concluded that “male” mentors and protegees had more benefits than “females”. The paper had to be retracted after receiving criticism about what counted as “informal mentorship”, and about the limitation of considering only the metrics of citation rates and impact to evaluate careers in science. The authors were accused of leaving aside important gender disparity issues like underrepresentation; the smaller access of women to grants and leadership roles, demonstrated in numerous studies and reports (18, 8, 19); and the salary gender gap present even in science (11). The authors “did not acknowledge their unjustified conclusions relating female gender to career success and policy suggestions” (20).

Publishers and academic associations have been forced to review their gender and racial biases, and publication and admission policies (21). Racism and sexual harassment events came to be addressed institutionally. In recent years, many journals and scientific associations have committed to reducing inequalities in publishing procedures and towards the equality of gender and the inclusion of diversities (22, 23, 19, 24, 25, among others). Bringing these situations to the public and producing data about the inequalities made publishing policies more explicit, and made inclusion and diversity into important topics to be considered for good scientific practices. But we still have a long way to go “to make significant changes for gender equality” (20).

Gender studies have contributed largely to legitimizing discussions about equity and representation in many areas of social life, such as science and technology. But “the question of science in feminism” (2) has also led to analyses that situate scientific discourse and practices as socially, culturally, and politically marked (26, 5, 27). In that sense, the very concept of “sex” or “sexual differences” became historically and culturally situated, shifting criteria for what counts as “female” and “male” from a given-natural-material-bodily difference to processes that are more open, variable, complex, and co-produced (28). The way each culture and society defines “female” and “male”, “women” and “men” vary greatly (29).

Menstruation and menstrual blood are tied within this complexity (30), being mostly associated to “female” bodies, bodies understood as feminine and of cis women – although trans men, non-binary people, women on menopause, women without uterus complicate this association. We cannot reduce the relations between menstruation and gender identities. In this study, we assume menstrual blood is a bodily fluid that socially and culturally marks differences between women and men, being understood as a specific phenomenon that happens to most cis women during a long period of their lives. We will consider its presence in scientific laboratories, relating to mesenchymal stromal/stem cells and to the scientists that work with it in their research. What can we tell about the relations between menstrual blood and gender when we look at research on menstrual blood mesenchymal stromal/stem cells (mbMSCs)? How are microscopes, scientific papers, journals, social networks, and power relations between women and men, senior and junior, global North and South articulated to co-produce differences related to gender and science? To address these questions, we will focus on results from research with mbMSC.

In the 1970’s, Friedenstein and colleagues were the first to report the existence of a type of adult stem cell in the bone marrow stroma, different from hematopoietic stem cells. These cells were clonogenic, adherent to the culture flask, of a fibroblastic shape (31, 32, 33), and named as mesenchymal stem cells (MSCs) (34). Since then, these cells have been extensively studied and their main mechanism of action has been described through paracrine secretion (35, 36). These cell-derived products, such as extracellular vesicles, trophic and immunomodulatory factors, have shown significant benefits in vivo(37, 38, 39, 40). Considering the growing publications demonstrating the beneficial paracrine effect promoted by the released factors, another proposed name for them was “Medicinal Signaling Cells” (41, 42).

The MSCs are found in a variety of tissues in the human body and in extra-embryonic attachments and are characterized according to the minimal criteria established by the International Society for Cell Therapy (43). However, the obtention of many of these MSCs needs invasive procedures, such as bone marrow and adipose MSCs. The MSC derived from extra-embryonic attachments are also only available once, at birth, or by invasive procedures during pregnancy, such as amniocentesis. In this context, menstrual blood is a unique MSC source, monthly available during all reproductive lives of cis women, and safely collected without invasive procedures by the donor. But we will demonstrate it is one of the least studied sources of MSCs despite all these advantages of working with mbMSC, such as its availability for decades throughout women’s lives and its easy and painless collection.

We analyzed the presence and prevalence of mbMSCs in scientific papers involving mesenchymal cells published in the past twelve years (2008 – 2020) and included in the Pubmed database. We compared the uses of mbMSCs with other mesenchymal stromal/stem cells also being studied (like bone marrow, umbilical cord, adipose tissue, placenta, dental pulp, amniotic fluid, and endometrium).

Our focus is on the relation between gender issues and the presence or absence of mbMSC in the group of bodily tissues usually employed in the fields of regenerative medicine, bioengineering, and cell therapy. How many of the papers published in the last twelve years about mesenchymal stromal/stem cells used menstrual blood as a source? Who were the scientists involved in these studies with menstrual blood in terms of gender? In which countries are their institutions located? Are they the same as mainstream publications in the field of stem cells? What is the impact factor of the journals that published research with mbMSCs? Do they look like the most important publications in the field? What is the social attention received by mbMSCs on Twitter? What could be improved in the communication of mbMSCs?

Our hypothesis is that menstrual blood is not a common source of mesenchymal stromal/stem cells, and that women scientists are responsible for most research using it. We also expect to see that research related to menstrual blood stem cells lacks social attention on Twitter, that the attention it gets is primarily by women, and that tweets may reveal bias against the use of mbMSC.

## Materials and Methods

Our analysis is focused on research about mesenchymal stromal/stem cells in the PubMed database. Pubmed is one of the most important archives of scientific literature in the fields of biomedical and life sciences. It is a publicly available repository for medical literature and probably the main source for medical literature (43) and, therefore, relevant to scientific research results on stem cells.

Considering the first studies about menstrual blood mesenchymal stromal/stem cells were published in 2007, we took 2008 as initial date for our search and applied a twelve-year period (2008 – 2020). The following terms and keywords were used: 2008 [Date – Publication]: 2020 [Date – Publication] AND human [Title/Abstract] OR mesenchymal [Title/Abstract] OR stromal [Title/Abstract] OR stem cell [Title/Abstract] AND menstrual blood [Title/Abstract]. A total of 229 results were found. All abstracts were individually read, and, in most of the publications, the item “materials and methods” too. Research that did not use menstrual blood as a source for mesenchymal cells was discarded. We have then selected 171 articles for the analysis.

The 171 articles were downloaded and tabulated with the following information: first author’s name, gender, country and university; last author’s name, gender, country and university; number of authors; journal in which the paper was published; year of publication; DOI; Pubmed ID; journal’s Impact Factor. The results allowed us to analyze the author’s profile, comparing gender, country and university, the journal’s Impact Factor, as well as the articles’ altmetrics in Twitter. We also mapped the flux of publications throughout the twelve years.

We considered “gender” as the social expression of a difference understood by western culture as “sexual”, and as something that can be represented by the person’s name and physical appearance. Genders of first and last authors were compared, along with the profiles of countries and universities. We aim at addressing the various scales or levels in which gender dynamics are involved: considering how women and men are present among these scientists and how cells, such as menstrual blood cells, come to be gendered and what that implies in terms of scientific results and impacts.

Gender assignment was made by inference using the first name and the profiles in universities’ websites, in academic social networks (academia.edu and researchgate.net), and in private social networks (Facebook and Twitter). Ambiguous names, especially of researchers from China and Japan, whose names do not have a gendered written form in Roman alphabet, were analyzed by using the software genderize.io. We have only considered the results with 60% or higher probability of being right. Cases of non-binary and trans people were disregarded in this analysis. Moreover, a chi-square test was performed using R software (version 4.1.1) to analyze the differences between the author’s genders. Table 1 shows the numbers for the three outcomes considered (female, male, and null) for the first and last author’s.

**Table 1.** Gender of first and last authors of studies about mbMSCs

|  | First Author | Last Author |
| --- | --- | --- |
| Female | 91 | 56 |
| Male | 69 | 109 |
| Null | 11 | 6 |

To compare the presence and prevalence of menstrual blood with other tissues of the body that also have mesenchymal cells, the same PubMed search was made with the following terms, as substitutions to “menstrual blood”: “bone marrow; umbilical cord; umbilical cord blood; umbilical cord vein; Wharton jelly; adipose; placenta; dental pulp; endometrium, and amniotic fluid”. The terms of search were: “2008” [Date – Publication]: “2020” [Date – Publication] AND human [Title/Abstract] OR mesenchymal [Title/Abstract] OR stromal [Title/Abstract] OR stem cell [Title/Abstract] AND the name of the cell’s origin [Title/Abstract].

As for the social attention analysis, we have used the DOIs, or PubMed ID of the publications sampled to track them on Twitter using the free API from Altmetric.com data provider. Altmetric.com is a data provider of social attention of science literature online. The AAS (altmetric attention score) measures how scientific publications (as articles, book chapters, preprints etc) were mentioned on online platforms such as blogs, news, Wikipedia, Twitter, and Facebook. We have used the DOIs or PubMed ID to track the 171 articles on Twitter using the free application programming interface (API) from Altmetric.com data provider.

We have manually collected 210 tweets (15.2%) mentioning 26 articles from a total of 352 tweets, since Altmetric.com provides only a limited free sample of tweets per paper for free. The tweets were then categorized by geolocation, profile gender (female, male, group, other or non-identified), profile type (science related, lay person, news, bot, non-identified) type of tweet (comment, retweet, or title and link of the paper), and ton of the comment (positive, negative, or neutral). The ton was considered neutral when it was not for or against the use of msMSC as most retweets or titles followed by the paper link; positive when optimistic, enthusiastic, or in favor of the use of msMSC, like sharing its benefits, treatments, or positive results in research; and negative when comments were demeaning, sarcastic, or sharing negative results or limitations in research.

## Results

### Research with menstrual blood cells is minoritarian and mostly conducted by women

**Table 2** shows the number of articles found for each of the tissues menstrual blood, bone marrow, umbilical cord, adipose, placenta, dental pulp, endometrium, and amniotic fluid. We considered endometrial cells as different from menstrual blood cells because they are usually obtained by invasive procedures such as hysterectomy, which might result in a different type of cell population. But since both endometrial and menstrual blood cells come from the uterus, we analyzed a sample of 45 papers mentioning endometrial cells (the 15 first, 15 last, and 15 in the middle of search results) to assure that they were not talking about menstrual blood cells when they mentioned endometrium. We read the abstracts and materials and methods of these papers, and only 2 of the 45 showed this ambiguity.

**Table 2.** Menstrual blood stromal/stem cells (mbMSCs) represent 0.25% of studies published at PubMed (2008 – 2020)

| Tissue | Number of results | Percentage |
| --- | --- | --- |
| menstrual blood | 229 | 0.25% |
| bone marrow | 43355 | 47.71% |
| umbilical cord total | 11013 | 12.12% |
| adipose | 20131 | 22.16% |
| placenta | 7721 | 8.50% |
| dental pulp | 3179 | 3.50% |
| endometrium | 3741 | 4.12% |
| amniotic fluid | 1488 | 1.64% |
| **Total** | **90857** | **100.00%** |

The comparative analysis shows an extremely low prevalence of publications about menstrual blood mesenchymal stromal/stem cells (0.25% of the research results for the period 2008-2020), in comparison with other tissues, such as bone marrow, umbilical cord, adipose, placenta, dental pulp, endometrium, and amniotic fluid.

Among the 171 articles published about mbMSCs selected from the initial 229, we have noticed a prevalence of women as first authors and men as last authors. We classified as “null” the authors whose gender identity we could not infer. **Table 1** and **Figure 1** show 53.2% [91] of first authors were women and 40.35% [69] were men, while men represented 63.74% [109] of last authors and women, 32.74% [56]. The non-identified first and last authors represent 6.43% [11] and 3.5% [6] of the studies, respectively. We established the statistical significance of these results regarding gender identity of the authors of mbMSC publications using a chi-square test (p = 0.00008303).

**Figure 1.**
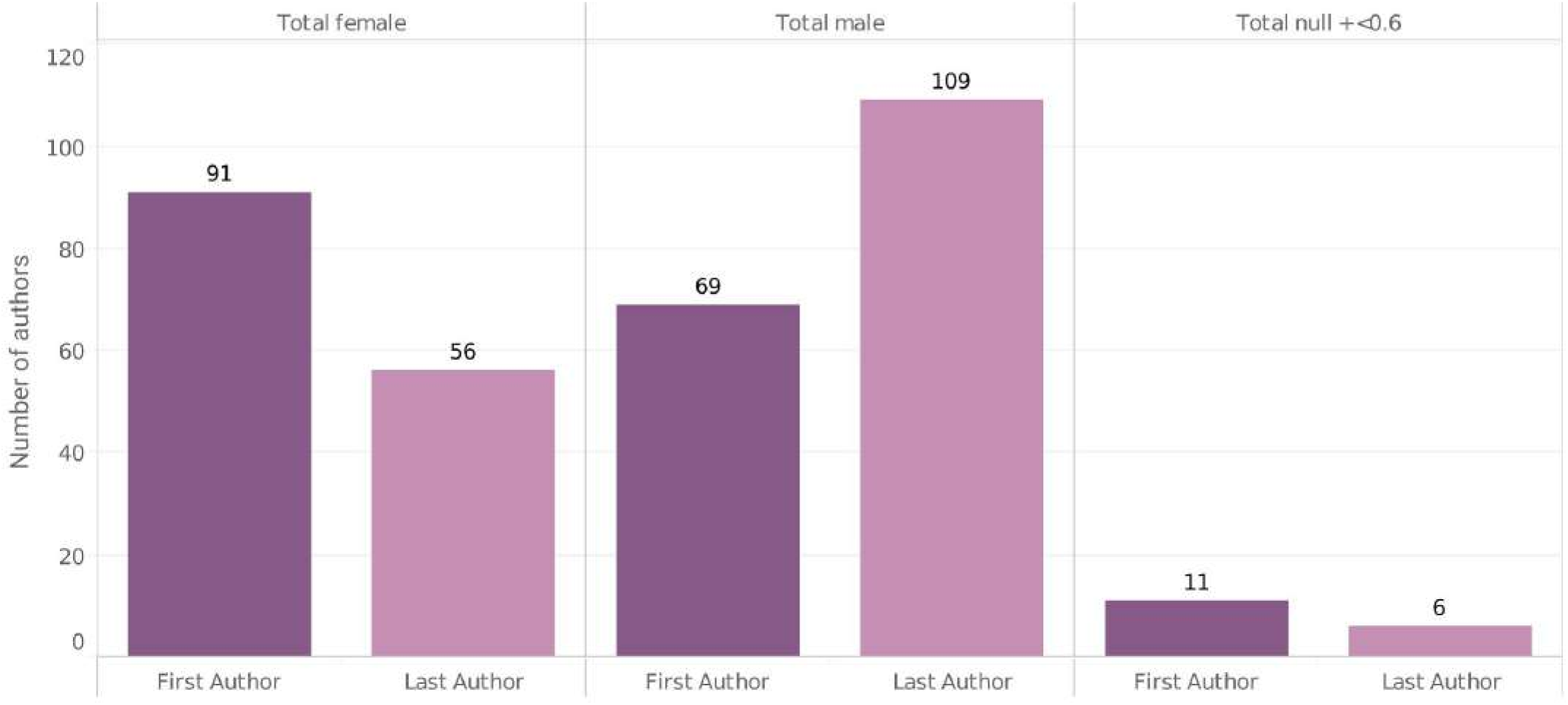
Gender of first and last authors of mbMSC publications.

Chinese scholars were the main authors (**Figure 2**), which represent 62 of the 171 articles analyzed (36.25%). The other authors come from institutions located mostly outside of Europe and the United States: Iran (40, 23.39%), Australia (8, 4.67%), and Brazil (7, 4.09%). The United States has 10 publications, but they come from private research centers (2, 1.17%), and from the University of South Florida (7, 4.09%) and Emory University (1, 0.58%). Spain (9) and the UK (5) represent 8.19% of the articles published.

**Figure 2.**
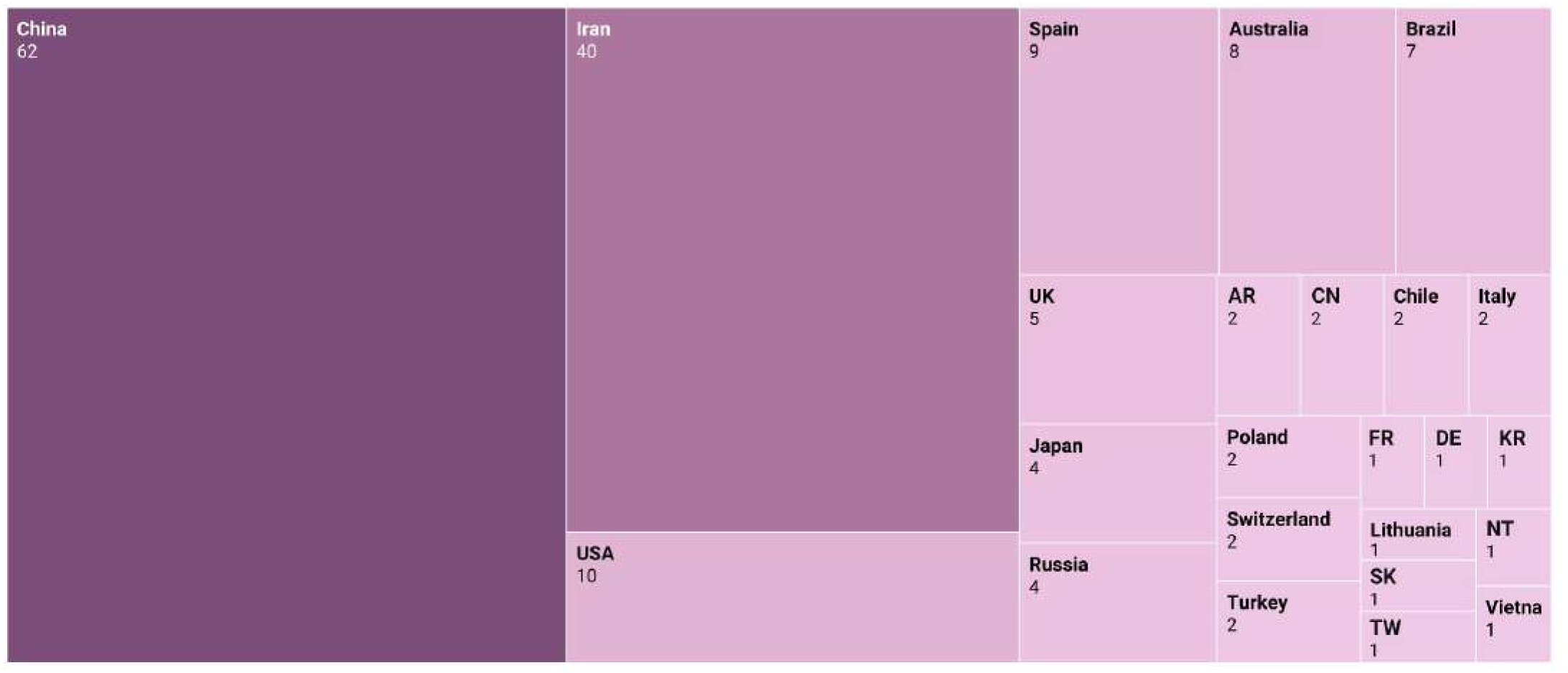
Country of the institution of origin of the last author.

**Figure 3** shows the Impact Factors attributed to the journals that have published the papers involving menstrual blood cells varied mostly between 1 and 5, suggesting that these publications tend to appear in journals with low/medium impact. Most journals (45.03%) have an impact factor between 3 and 4.99.

**Figure 3.**
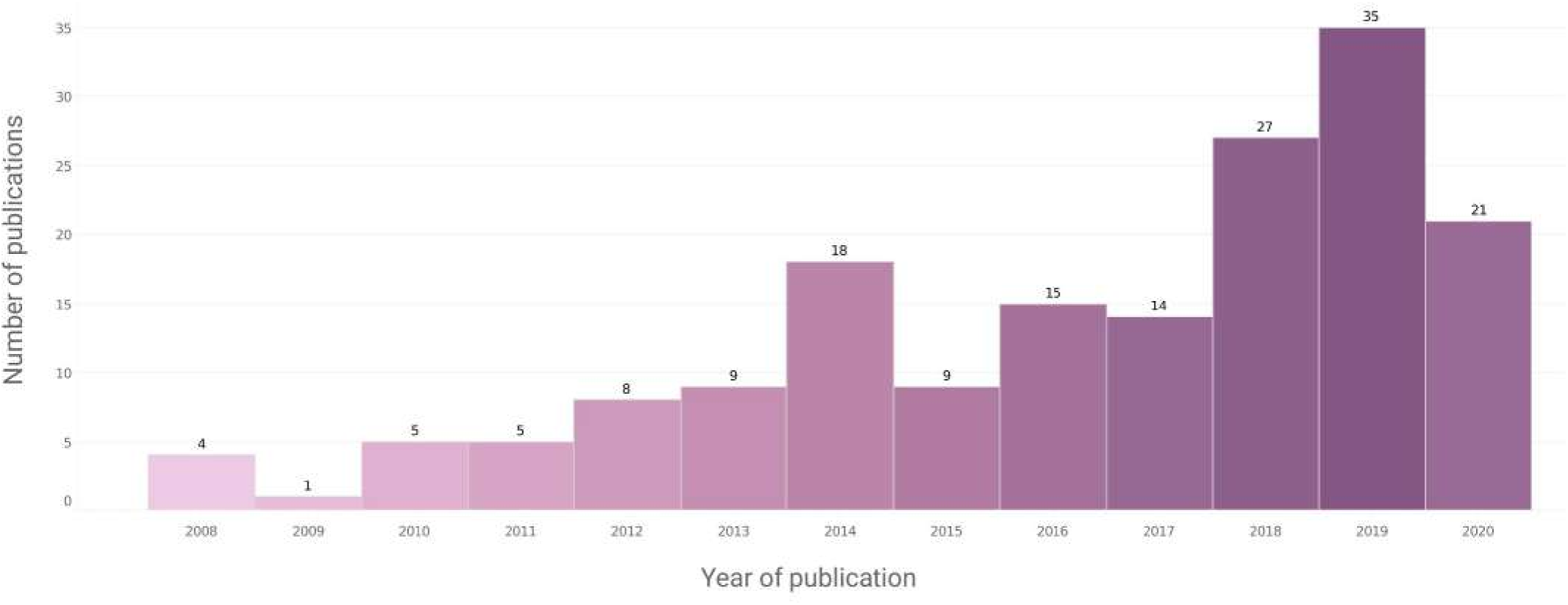
Number of publications with mbMSCs per year.

**Figure 4** shows a trend of increasing number of publications, with most papers (83, 48.53%) published since 2018, indicating a growing potential for research involving menstrual blood mesenchymal stromal/stem cells, despite its still small participation among the most representative tissues in scientific research and medical therapies with mesenchymal cells.

**Figure 4.**
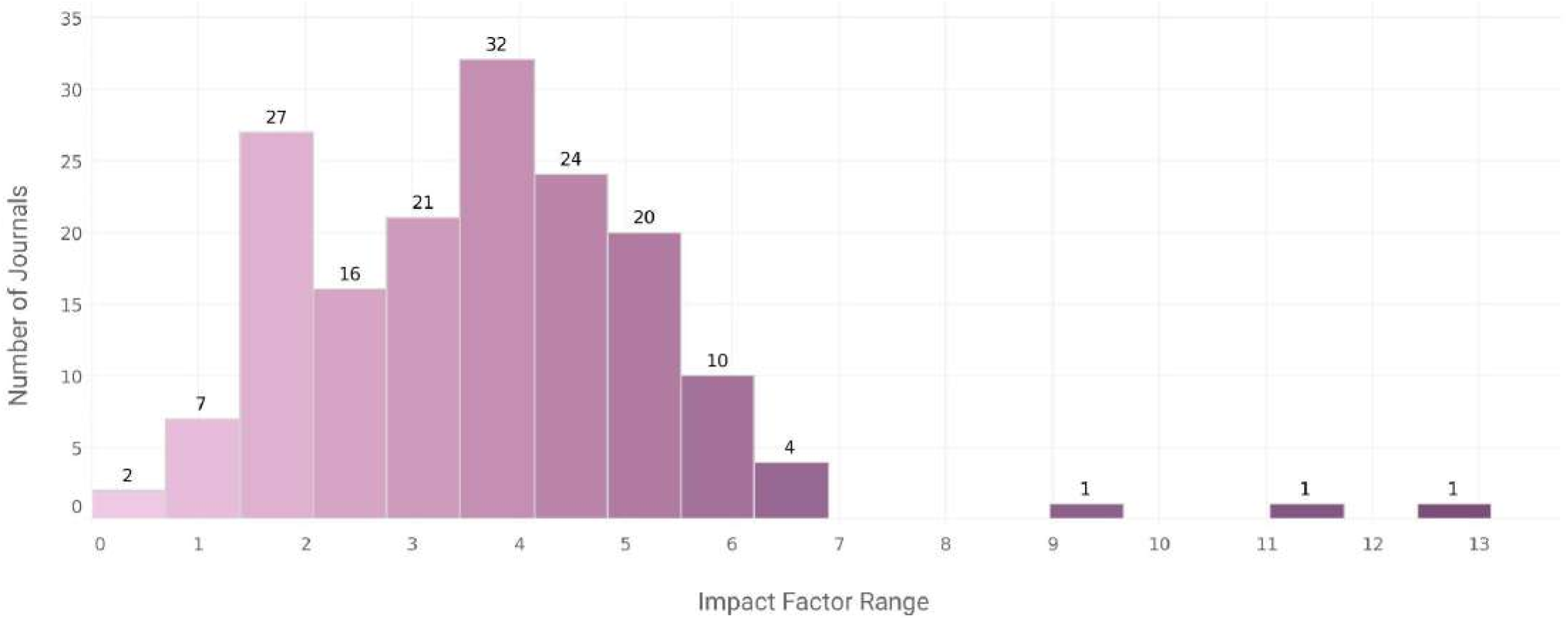
Impact factors of journals that published mbMSCs papers.

### Social attention of mbMSC on Twitter

Within our sample, 40.9% of the papers received no social attention (AAS), which is a smaller number than observed for other publications (45), indicating social interest in mbMSC. Yet, the average AAS was 3.0 per paper, which is lower than the score found in other analyses (idem). Only 8.2% got an altmetric score higher than 10, all of which published in open or free access papers, an advantage already reported on altmetrics research (46).

The analyses of these tweets showed that 35.5% of them were comments, which are the best type of engagement when compared to retweets (32.7%) or to simply sharing the paper title and link (31.8%). Most comments were positive about mbMSC (77.6%), neutral (20.8%), or negative (11.7%), like what stem cell tweet analysis showed (47, 48), and most of them were from lay people. Women with Twitter profiles related to civil rights or feminism made most of the positive comments (36.4%), followed by groups (15.6%) – many of which were surprisingly related to investments or stock market (8.1% among all tweets) –, and men (13%) – mostly scientists. Groups (news feed, labs, institutions, bots, for example) published most of the tweets (38.1%), but they were mainly retweets or just sharing paper titles and links. Therefore, these tweets’ goal is probably to disseminate new papers. All negative comments were made by men (6 profiles mainly made by non-scientists), mostly questioning the plausibility and sterility of using these cells [6 comments were related to the article “The Potential of Menstrual Blood-Derived Stem Cells in Differentiation to Epidermal Lineage: A Preliminary Report” (**Supporting Information Table 1**)]. But men also shared some positive (13%) or neutral (7.8%) comments.

Around 61.9% of the tweets were geolocated, a value close to what Robillard et al. (2015) (48) found in studies about stem cells (from 63% to 65%). The most active countries were the USA (37.7%), Australia (7.7%), the UK (6.9%), Japan (6.1%), and Mexico (6.1%), which were different countries from the ones our results indicated as more engaged with mbMSC research.

According to the profile of tweet authorship, most (33.8%) were science related (scientists, professors, PhD students, doctor, institutions, journals, and so on), but with intense exchange with lay people (25.7%), investment groups (8%), news feeds (6.2%), bots (5.2%), and others (9%).

## Discussion

### Menstrual Blood Derived Mesenchymal Stem Cells: State of the Art

Studies published using mbMSCs or its products indicate a promising therapeutic potential for several disease models, showing significant benefits in vitroand in vivo. These cells have already been transplanted intracerebrally and intravenously in a rat model of ischemic stroke and reduced behavioral and histological disorders (49). In a mouse liver fibrosis model, mbMSC migrated to the injury site, improved liver function, inhibited activated hepatic stellate cells and reduced collagen deposition (50). The therapeutic capacity of mbMSC has also been tested in a pulmonary fibrosis mouse model. In thisstudy, cells migrated to the lung and improved its structure, reducing collagen deposition and inflammatory response (51). The mbMSC also improved mouse embryonic development in vitro, until the blastocyst stage (52). Additionally, mbMSC showed good results for in vivomodels of myocardial infarction (53, 54), acute liver injury (55), spinal cord injury (56), duchenne muscular dystrophy (57), limb ischemia (58), wound healing (59, 60), and female reproductive system disorders (61, 62, 63, 64).

However, despite these positive results, the accessibility of the cell, and its abundant availability, obtained by painless and non-invasive methods, mbMSCs are not among the most studied sources in non-clinical and clinical studies. Until the year of 2020, only three clinical trials were registered in the clinicaltrials.gov database using mbMSCs (NCT01496339, NCT01483248, NCT01558908). Given the absence of technical explanations for that small number, we would like to suggest a gender bias concerning the choices of MSCs sources. In other words, menstrual blood’s relation to gender, as a “feminine” bodily tissue, could explain mbMSC’s lack of expressivity in MSC research.

### Women in benchwork, men as lab chiefs

In Biological Sciences, first authors are usually researchers directly involved with benchwork and with most of the work of writing the paper and organizing the results, while last authors tend to be senior academics, the principal investigators, and tenured professors. Women are underrepresented among last authors, compared to men (15), who figure as most researchers and laboratory supervisors. Our results found that men are more prevalent as last authors, confirming the tendency of having less women among senior researchers, and occupying the higher hierarchical academic positions (18, 8).

Our results also showed that menstrual blood research is mostly conducted by women scientists, which is different from what other reports on gender and science have demonstrated. As West (2013) (15) argues, for example, men usually predominate as first authors in most fields. This result suggests women may be more willing to work with menstrual blood than men. And, as our qualitative research has shown, in contexts in which access to cells and research materials is restricted, women scientists often provide the bodily material themselves, and they tend to have a greater ease to work with menstrual blood, despite jokes and demonstrations of disgust from other scientists (65). Concerns found in social media in commentaries made by men about the “sterility” of menstrual blood as a source for cells also confirm a well-known association between menstruation and dirt, impurity, and pollution in western culture (66).

### Publications from the margins

Europe and the US have produced most of the research about stem cells between 2000 – 2010 (67). Our results show that the papers on menstrual blood cells were mainly published by Chinese scholars, situated in Chinese universities. Other research centers are in countries (Iran, Australia, and Brazil) less present among those that concentrate the production on stem cells and in mesenchymal stem cells (67, 68). The prevalent institutions in the field are Harvard University, University of California, Johns Hopkins, and Stanford (68), but none of them figure among the American institutions that published results with mbMSCs. This suggests that menstrual blood is unprivileged by the most renowned universities, and that the institutions that have been studying the potentials of mbMSC are less traditional in the stem cell research field.

Analysis of the journals’ impact factors include menstrual blood research within the array of a low/regular-impact scientific production, which confirms mbMSC’s absence in higher impact scientific journals (69, 68). Most papers were published in the last three years, which demonstrates a growing potential for menstrual blood mesenchymal stromal/stem cells research, despite the obstacles to its inclusion among the most representative tissues in scientific research and medical therapies with mesenchymal cells.

### Menstrual blood cells research on Twitter: potentially interesting

Studies have shown that women’s scientific work is less cited and gets less visibility on social media than men’s (70). Social media is an important space for gaining credibility, visibility, and success, therefore, relevant to strengthen the presence of women in science. Vásárhelyi and colleagues have concluded that even in fields where women are better represented, such as medical sciences, their online presence on social media remains lower (39%) than men’s (61%).

Communication on social media has grown worldwide and became a key space to monitor social interaction with information. To measure the impact of science on society, altmetrics emerge as relevant complementary metrics to measure the social attention that scholarly literature receives on online platforms (71). It is a faster and more diverse way to measure the impact of science than the traditional metrics. Yet, the social attention that science outputs receive on social media may differ from that of scholarly interest (72).

Around 49% of the world population use social media, 397,000 people have a Twitter account (73), among which women correspond to 31.9% (74). Twitter has become one of the main social medias to track social attention scores (such as the Altmetric Attention Score, AAS) and it is widely used among scholars (75, 76).

Our analysis has confirmed that women are more engaged with the social debate related to menstrual blood stem cells, and that men, despite also contributing with positive comments, are responsible for all the negative tweet comments. This could suggest that women researchers of mbMSC should be more active on Twitter to reinforce the visibility of their work and of their research field.

We found that scientists, institutions, and journals responsible for merely sharing paper titles and links could try to engage the audience in online debates and interactions, and this is required for increased visibility according to social media mechanics. Tweets about mbMSC papers tend to get social attention and interest, and therefore have great potential to communicate scientific results broadly, as suggested by the good number of tweets from lay people, including feminist and profiles related to investment or stock market.

Contrary to our hypothesis, tweets reveal no bias against the use of mbMSC, since positive comments were almost six times more frequent than negative comments.

Gender differences cannot be seen through the microscope, and despite mbMSCs attending all the required expectancies regarding the potential of mesenchymal stromal/stem cells, some work still needs to be done for menstrual blood to occupy its proper place in the universe of mesenchymal cell research.

### Limitations of the study

Pubmed database presented instabilities and changes in the online platform during our research. Some papers appeared simultaneously in two years, especially when approved and published in different years. We have identified and excluded them from the table.

Inferring gender by names disregards many important variables, like gender transition, non-binary or androgynous names or physical appearances. Gender cannot often be inferred for Chinese names in the Roman alphabet. Nonetheless, we have considered it was still important to look for differences between what could be considered as men and women among first and last authors, using the software genderize.io to help us infer gender in those cases, and others.

As for the analysis with altmetrics.com, we know the tweets analyzed cannot be generalized. Yet, our sample corresponds to 55.5% of the tweets received by the selected mbMSCs article and considering studies with more tweets and higher AAS, to select the most representative expressions about the studies.

## Supporting information

Suppl Inf1_Table with research output at Pubmed

## Data Availability

All relevant data are within the manuscript and its Supporting Information files, and they can also be found in .ods format at the REDU/Unicamp database: https://doi.org/10.25824/redu/NKJV2D

## Acknowledgments

This research is funded by Fapesp (SP/Brazil), process number 2018/20651-3. We would like to thank Asura Enkhbayar, from #ScholCommLab at Simon Fraser University for providing Altmetrics data. We would also like to thank Bernardo Tura, from National Institute of Cardiology, Research and Teaching Department Rio de Janeiro, RJ, Brazil, for providing statistical results. We thank Pedro Ferreira for a first review of the manuscript, and Clarissa Reche for a support with the graphics. The authors thank Espaço da Escrita (Pró-Reitoria de Pesquisa – UNICAMP) for the language services provided.

## Author contributions

**Conceptualization**: Daniela Manica; Germana Barata

**Data Curation**: Daniela Manica; Gaia Mazzarelli; Karina Asensi

**Formal Analysis:** Karina Asensi; Germana Barata; Regina Goldenberg

**Funding Acquisition:** Daniela Manica

**Investigation:** Gaia Mazzarelli; Germana Barata

**Methodology:** Daniela Manica; Germana Barata

**Project Administration:** Daniela Manica

**Resources:** Daniela Manica

**Supervision:** Daniela Manica

**Visualization:** Gaia Mazarelli

**Writing - Original Draft:** Daniela Manica; Karina Asensi

**Writing - Review & Editing:** Daniela Manica; Karina Asensi; Germana Barata; Regina Goldenberg

