## Supplementary material for "Can we see gender through the microscope? The research output using menstrual blood cells has much to say": Suppl Inf1_Table with research output at Pubmed

| First author | Sex | Sex-Software | Country (1st author) | Last author | Sex | Sex-Softw | Country (last author) | Publishing date | Journal - title | Impact Factor | DOI |
|---|---|---|---|---|---|---|---|---|---|---|---|
| Caroline Gargett | F | | Australia | James Deane | M | | Australia | 2015 | Human Reproduction Update | 11.194 | 10.1093/humupd/dmv051 |
| Maria Carolina Rodrigues | F | | Brazil | Cesar Borlongan | M | | USA | 2016 | Biobanking and Cryopreservation of Stem Cells (B | book | 10.1007/978-3-319-45457-3 |
| Connor Stonesifer | M | | USA | Cesar Borlongan | M | | USA | 2017 | Progress in Neurobiology | 12.716 | 10.1016/j.pneurobio.2017.07.004 |
| Lu Chen | | F 0.57 | China | Charlie Xiang | M | | China | 2017 | Stem Cell Research & Therapy | 5.278 | 10.1186/s13287-016-0453-6 |
| Cesar Borlongan | M | | USA | Paul R Sanberg | M | | USA | 2010 | Stem Cells And Development | 4.922 | 10.1089/scd.2009.0340 |
| Fahimeh Tabatabaei | F | | Iran | Jafah Ai | M | | Iran | 2017 | Regenerative Medicine | 2.664 | 10.2217/rme-2017-0029 |
| Ilona Uzielienė | F | | Lithuania | Eiva Bernotiene | F | | Lithuania | 2018 | Stem Cells International | 3.937 | 10.1155/2018/5748126 |
| Huaijuan Ren | | null | China | Yantian Chen | | F 1.0 | China | 2015 | Stem Cells International | 4.616 | 10.1155/2016/3516574 |
| Yanli Liu | | F 0.87 | China | Juntang Lin | | M 1.0 | China | 2017 | Journal of Cellular and Molecular Medicine | 4.439 | 10.1111/jcmm.13437 |
| Bingyu Xiang | | F 0.64 | China | Charlie Xiang | M | | China | 2017 | International Journal of Molecular Sciences | 3.962 | 10.3390/ijms18040689 |
| David Eve | M | | USA | Krystyna Domanska | F | | Poland | 2018 | Human Neural Stem Cells (Book) | book | 10.1007/978-3-319-93485-3 |
| Martin A Ivarsson | M | | UK | Niklas K. Bjorkstrom | M | | UK | 2018 | Mucosal Immunology | 6.859 | 10.1038/mi.2016.50 |
| Hossein Eyni | M | | Iran | Masoud Soleimani | M | | Iran | 2017 | Journal of Biomaterial Applications | 2.264 | 10.1177/0885328217723179 |
| Dongmei Lai | | F 0.98 | China | Charlie Xiang | M | | China | 2015 | Journal of Translational Medicine | 4.222 | 10.1186/s12967-015-0516-y |
| Haining Lv | | M 1.0 | China | Huidong Jia | M | | UK | 2018 | Stem Cell Research & Therapy | 4.777 | 10.1186/s13287-018-1067-y |
| Mehdi Aleahmad | M | | Iran | Siwen Zhang | M | | Iran | 2018 | Avicenna Journal of Medical Biotechnology | 1.55 | PMID: 30090214 |
| Xiao-Jun Wang | M | | China | Charlie Xiang | M | | China | 2017 | Oncotarget | 4.849 | 10.18632/oncotarget.17621 |
| Saeed Farzamfar | M | | Iran | Majid Salehi | M | | Iran | 2017 | Molecular Biology Reports | 2.178 | 10.1007/s11033-017-4124-1 |
| Zahra Rajabi | F | | Iran | Amir-Hassan Zarnar | M | | Iran | 2018 | Reproductive Biology | 2.043 | 10.1016/j.repbio.2018.02.001 |
| Karina Asensi | F | | Brazil | Regina Goldenberg | F | | Brazil | 2014 | Journal of Cellular and Molecular Medicine | 4.605 | 10.1111/jcmm.12226 |
| Razieh Dalirfardouei | F | | Iran | Elahe Mahdipour | F | | Iran | 2018 | Tissue and Cell | 1.553 | 10.1016/j.tice.2018.09.010 |
| Yanling Zhang | F | | China | Songying Zhang | F | | China | 2016 | Reproduction Research | 3.514 | 10.1530/REP-16-0286 |
| Nikoo Shojanoori | F | | Iran | Amir-Hassan Zarnar | M | | Iran | 2012 | The Journal of Obstetrics and Gynaecology Resea | 1.128 | 10.1111/j.1447-0756.2011.01800.x |
| Shulan Zhang | F | | China | Jingwei Tan | F | | China | 2018 | Stem Cell Research & Therapy | 4.777 | 10.1186/s13287-018-0795-3 |
| Fereshteh Azedi | F | | Iran | Shaghayegh Arastel | F | | Iran | 2017 | Molecular Biology Reports | 2.178 | 10.1007/s11033-016-4095-7 |
| Dongmei Lai | | F 0.98 | China | Charlie Xiang | M | | China | 2016 | Acta Biochimica et Biophysica Sinica | 2.148 | 10.1093/abbs/gmw090 |
| Yuliang Sun | | M 0.93 | China | Juntang Lin | | M 1.0 | China | 2019 | Biology Open | 2.026 | 10.1242/bio.038885 |
| Paul R. Sanberg | M | | USA | Cesar V Borlongan | M | | USA | 2011 | Cell Transplantation | 4.116 | 10.3727/096368910X532855 |
| Daniela Ulrich | F | | Australia | Caroline Gargett | F | | Australia | 2013 | Expert Opinion Biological Therapy | 4.120 | 10.1517/14712598.2013.826187 |
| Kana Sugawara | | F 0.8 | Japan | Akihiro Umezawa | M | | Japan | 2014 | Scientific Reports | 5.990 | 10.1038/srep04599 |
| Mahmood Bozorgmerh | M | | Iran | Amir-Hassan Zarnar | M | | Iran | 2014 | Immunology Letters | 2.868 | 10.1016/j.imlet.2014.10.005 |
| Mina Fathi-Kazerooni | F | | Iran | Somaieh Kazemneja | F | | Iran | 2017 | Cytotherapy | 4.017 | 10.1016/j.jcyt.2017.08.022 |
| Davood Mehrabani | M | | Iran | Farnaz Ghobadi | F | | Iran | 2016 | Iranian Journal of Medical Sciences | 0.94 | PMID: 26989284 |
| Filippo Rossignoli | M | | Italy | Massimo Dominici | M | | Italy | 2013 | BioMed Research International | 3.276 | 10.1155/2013/901821 |
| Shohreh Nikoo | F | | Iran | Amir-Hassan Zarnar | M | | Iran | 2014 | Molecular Human Reproduction | 4.209 | 10.1093/molehr/gau044 |
| Frederico Allison-Silva | M | | Brazil | Adriane Todeschini | F | | Brazil | 2014 | Glycobiology | 3.595 | 10.1093/glycob/cwu012 |
| Yanping Xu | | F 0.7 | China | Jichun Tan | M | | China | 2015 | International Journal of Clinical and Experimental | 1.316 | PMID: 26885178 |
| Xue Du | F | | China | Jia Bei | F | | UK | 2016 | Stem Cells International | 3.756 | 10.1155/2016/3573846 |
| R Moreno | | | Spain | Rafael Alemany | M | | Spain | 2017 | Stem Cells International | 3.959 | 10.1155/2017/3615729 |
| Te Liu | M | | China | Chuan Chen | | M 0.86 | China | 2014 | Stem Cells International | 3.582 | 10.1089/scd.2013.0371 |
| Marina V. Kovina | F | | Russia | Alexey V. Lyubdup | M | | Russia | 2018 | Cytotherapy | 4.120 | 10.1016/j.jcyt.2017.12.012 |
| Danubia Silva dos Santos | F | | Brazil | Regina Goldenberg | F | | Brazil | 2014 | Cell Medicine | | 10.3727/215517914X679265 |
| Saeedeh Darzi | F | | Iran | Somaieh Kazemneja | F | | Iran | 2012 | Tissue Engineering | 4.946 | 10.1089/ten.tea.2011.0386 |
| Irina Kozhkharova | F | | Russia | Nikolay Nikolsky | M | | Russia | 2017 | International Journal of Hematology | 2.152 | 10.1007/s12185-017-2346-6 |
| Lijun Chen | | F 0.51 | China | Charlie Xiang | M | | China | 2016 | Stem Cells Translational Medicine | 4.672 | 10.5966/sctm.2015-0265 |
| Natalia Russo | F | | Italy | Andrea Riccardo Ge | M | | Italy | 2012 | Gynecological Endocrinology | 1.607 | 10.3109/09513590.2011.633667 |
| Rafael Moreno | M | | Spain | Ramon Alemany | M | | Spain | 2018 | Molecular Cancer Therapeutics | 5.151 | 10.1158/1535-7163.MCT-18-0431 |
| Deivid de Carvalho Rodri | M | | Brazil | Turán Péter Ürmény | M | | Brazil | 2012 | Cell Transplantation | 4.810 | 10.3727/096368912X653048 |
| Saeed Farzamfar | M | | Iran | Mehdi Aleahmad | M | | Iran | 2018 | Biomedical Engineering Letters | 1,2 | 10.1007/s13534-018-0084-1 |
| Naoko Hida | F | | Japan | Akihiro Umezawa | M | | Japan | 2008 | Stem Cells | 9.162 | 10.1634/stemcells.2007-0826 |
| Yongjia Zhao | | M 0.71 | Japan | Charlie Xiang | M | | Japan | 2018 | Frontiers in Molecular Neuroscience | 3.814 | 10.3389/fnmol.2018.00140 |
| Zhaocai Zhang | | M 1.0 | China | Hong Yo | | M 0.65 | China | 2013 | International Journal of Cardiology | 1.882 | 10.1016/j.ijcard.2013.03.126 |

| | | | | | | | | | | | |
|---|---|---|---|---|---|---|---|---|---|---|---|
| Patricia Luz-Crawford | F | | Chile | Maroun Khoury | M | | Chile | 2015 | Stem Cells | 6.585 | 10.1002/stem.2244 |
| Dah-Ching Ding | F | | Taiwan | Shinn-Zong Lin | M | | Taiwan | 2011 | Cell Transplatation | 4.116 | 10.3727/096368910X |
| Jinyang Chen | | M 0.69 | China | Charlie Xiang | M | | China | 2015 | International Journal of Clinical and Experimental | 1.825 | PMID: 26823782 |
| Jichun Tan | M | | China | Lin Kong | | F 0.63 | China | 2016 | Human Reproducction | 5.020 | 10.1093/humrep/dew235 |
| Julie Allickson | F | | USA | Paul R. Sanberg | M | | USA | 2011 | Open Stem Cell Journal | | 10.2174/1876893801103010004 |
| Haiyan Zhu | | F 0.9 | China | Songying Zhang | F | | China | 2018 | Reproductive Biology | 2.043 | 10.1016/j.repbio.2018.06.003 |
| Birol Ay | M | | Turkey | Halime Kenar | F | | Turkey | 2016 | Journal of Biomedical Materials Research Part A | 3.607 | 10.1002/jbm.a.35948 |
| Jian Lin | | M 0.9 | China | Charlie Xiang | M | | China | 2011 | Journal of Zhejiang University | 1.482 | 10.1631/jzus.B1100015 |
| Shanti Gurung | F | | Australia | Caroline E. Garget | F | | Australia | 2015 | Seminars in Reproductive Medicine | 2.776 | 10.1055/s-0035-1558405 |
| Shaghayegh Arasteh | F | | Iran | **Somaieh Kazemne** | F | | Iran | 2018 | Stem Cell Nanotechnology (book) | book | 10.1007/7651_2018_193 |
| Maroun Khoury | M | | Chile | Fernando E. Figuero | M | | Chile | 2014 | Frontier in Immunology | 4.994 | 10.3389/fimmu.2014.00205 |
| Shixia Bu | | F 1.0 | China | Dongmei Lai | | F 0.98 | China | 2016 | Scientific Reports | 4.738 | 10.1038/srep37019 |
| Ozge Karadas | F | | Turkey | Vasif Hasirci | M | | Turkey | 2012 | Journal of Tissue Engeneering and Regenerative | 2.989 | 10.1002/term.1555 |
| Somaieh Kazemnejad | F | | Iran | Saghari S | M | | Iran | 2013 | Journal of Stem Cells and Regenerative Medicine | 0.22 | 10.46582/jsrm.0901004 |
| Caroline E. Gargett | F | | Australia | Hirotaka Masuda | M | | Australia | 2010 | Molecular Human Reproducction | 3.660 | 10.1093/molehr/gaq061 |
| Sheng-Xia Zheng | | M 1.0 | China | Yu-Sheng Liu | M | | China | 2018 | International Journal of Molecular Medicine | 3.024 | 10.3892/ijmm.2018.3415 |
| Xuqi Hu | | F 1.0 | China | Huazi Xu | | M 0.64 | China | 2014 | Spine Journal | 2.445 | 10.1097/BRS.0000000000000261 |
| Hossein Faramarzi | M | | Iran | Reza Shirazi | M | | Iran | 2016 | World Journal of Plastic Surgery | | PMID: 27308237 |
| Qinfeng Wu | | M 1.0 | China | Chuanming Dong | | M 1.0 | China | 2018 | Cell Death and Disease | 6.083 | 10.1038/s41419-018-0847-8 |
| Julie Allickson | F | | USA | Charlie Xiang | M | | China | 2012 | Journal of Zhejiang University - Science B | 1.456 | 10.1631/jzus.B1200062 |
| Yijing Zheng | | F 0.88 | China | Jianjun Hong | | M 0.97 | China | 2017 | Experimental and Therapeutic Medicine | 1.551 | 10.3892/etm.2017.4383 |
| Manijeh Khanmohammad | F | | Iran | Somaieh Kazemneja | F | | Iran | 2012 | International Journal of Hematology | 2.106 | 10.1007/s12185-012-1067-0 |
| Xiao- Zhou Mou | M (foto | M 0.75 | China | Charlie Xiang | M | | China | 2013 | Journal of Zhejiang University - Science B | 1.733 | 10.1631/jzus.B1300081 |
| Stanimir Kyurkchiev | M | | Bulgaria | Rumen Dimitrov | M | | Bulgaria | 2010 | Reproducttive BioMedicine Online | 2.869 | 10.1016/j.rbmo.2009.12.011 |
| Maryam Fard | F | | Iran | Reza Shirazi | M | | Iran | 2018 | Molecular Biotechnology | 1.811 | 10.1007/s12033-017-0049-0 |
| Haitao Ren | | M 0.94 | China | Ruolang Pan | | null | China | 2018 | Stem Cells International | 3.937 | 10.1155/2018/7873625 |
| Xiaoxi Xu | | F 0.6 | Canada | Hao Wang | M | | Canada | 2016 | Stem Cells Translational Medicine | 4.672 | 10.5966/sctm.2016-0206 |
| Yanli Liu | | F 0.87 | China | Juntang Lin | | F 0.98 | China | 2018 | Stem Cells International | 3.937 | 10.1155/2018/3250379 |
| Naoki Tajiri | M | | USA | Yuji Kaneko | M | | USA | 2014 | International Journal of Molecular Sciences | 3.385 | 10.3390/ijms150915225 |
| Amit N. Patel | M | | USA | Julie allickson | F | | USA | 2008 | Cell Transplantation | 4.356 | 10.3727/096368908784153922 |
| Manijeh Khanmohammad | F | | Iran | Somaieh Kazemneja | F | | Iran | 2014 | Cell Proliferation | 3.734 | 10.1111/cpr.12133 |
| Ana Laura Alfano | F | | Argentina | Veronica Lopez | F | | Argentina | 2017 | Molecular Therapy Oncolutics | 3.915 | 10.1016/j.omto.2017.06.002 |
| Alisa P. Domnina | F | | Russia | Nikolay N. Nikolsky | | M 0.99 | Russia | 2016 | Experimental and Therapeutic Medicne | 1.490 | 10.3892/etm.2016.3671 |
| Bruna R. Sousa | F | | Brazil | Rodrigo R. Resende | M | | Brazil | 2013 | Cytometry Part A | 3.421 | 10.1002/cyto.a.22402 |
| Sayeh Khanjani | F | | Iran | Somaieh Kazemneja | F | | Iran | 2014 | PLOS ONE | 3.778 | 10.1371/journal.pone.0086075 |
| Xiaohan Wang | | F 0.69 | China | Cuifang Hao | | null | China | 2018 | Journal of Cellular Biochemistry | 3.448 | 10.1002/jcb.28014 |
| JIn-Yang Chen | | M 0.69 | China | Charlie Xiang | M | | China | 2015 | Asian Pacific Journal of Tropical Medicine | 1.630 | 10.1016/j.apjtm.2015.07.022 |
| Yang Li | | M 0.64 | China | Jichun Tan | M | | China | 2013 | Stem Cells and Development | 4.710 | 10.1089/scd.2012.0428 |
| Pham Van Phuc | M | | Vietnam | Phan Kim Ngoc | M | | Vietnam | 2011 | In Vitro Cellular & Developmental Biology - Anima | 1.665 | 10.1007/s11626-011-9399-2 |
| Federica Marino | F | | Spain | Beatriz Macías- Gar | F | | Spain | 2018 | Reproduction in Domestic Animals | 1.614 | 10.1111/rda.13314 |
| Somaieh Kazemnejad | F | | Iran | Kamran Alimoghada | M | | Iran | 2012 | The International Journal of Artificial Organs | 1.961 | 10.5301/ijao.5000019 |
| Sayeh Khanjani | F | | Iran | Somaieh Kazemneja | F | | Iran | 2015 | Journal of Tissue Engineering and Regenerative M | 3.211 | 10.1002/term.1715 |
| Xiaoxing Wu | | M 0.56 | China | Charlie Xiang | M | | China | 2014 | Stem Cells and Development | 4.349 | 10.1089/scd.2013.0390 |
| Marjan D. Manshadi | F | | Iran | Mehdi Abbasi | M | | Iran | 2018 | Microscopy Research and Technique | 1.355 | 10.1002/jemt.23120 |
| Javad Verdi | M | | UK | Alexander M. Seifali | M | | UK | 2014 | Journal of Biological Engeneering | 3.143 | 10.1186/1754-1611-8-20 |
| Fengyi Gou | | M 0.89 | China | Xue Du | F | | China | 2019 | Reproductive Biology and Endocrionology | 3.498 | 10.1186/s12958-019-0499-2 |
| Zhi Jaing | | M 0.75 | China | Jian-an Wang | M | | China | 2013 | Journal of Cellular and Molecular Medicine | 4.258 | 10.1111/jcmm.12100 |
| Fereshteh Azedi | F | | Iran | Niknam Lakpour | M | | Iran | 2014 | Cell Biollogy International | 2.160 | 10.1002/cbin.10245 |
| Yongcheng Lv | | | China | Hao Wang | | M 0.89 | China | 2014 | Journal of Translational Medicine | 4.630 | 10.1186/s12967-014-0344-5 |
| Amit N. Patel | M | | USA | Francisco Silva | M | | Argentina | 2008 | Regenerative Medicine | 2.950 | 10.1016/j.hfc.2014.12.006 |
| Maryam Rahimi | F | | Iran | Somaieh Kazemneja | F | | Iran | 2014 | Molecular Biotechnology | 2.215 | 10.1007/s12033-014-9795-4 |
| Shanzheng Lu | | | China | Hao Wang | | M 0.89 | China | 2016 | Journal of Translational Medicine | 4.155 | 10.1186/s12967-016-1051-1 |
| Hongyun Huang | | M 1.0 | China | Paul Sanberg | M | | USA | 2010 | Cell Medicine | | 10.3727/215517910X516673 |

| | | | | | | | | | | | |
|---|---|---|---|---|---|---|---|---|---|---|---|
| James A. Deane | M | | Australia | Caroline E. Gargett | F | | Australia | 2013 | Current Opinion in Obstetrics & Gynecology | 3.061 | 10.1097/GCO.0b013e32836024e7 |
| Yukinori Ikegami | M | | Japan | Akihiro Umezawa | M | | Japan | 2010 | Artificial Organs | 1.654 | 10.1111/j.1525-1594.2009.00859.x |
| Jia Hu | | F 0.63 | China | Bu-Zhen Tan | | Null | China | 2019 | Molecular Medicine Reports | 2.085 | 10.3892/mmr.2018.9744 |
| Maryam Rahimi | F | | Iran | Somaieh Kazemneja | F | | Iran | 2014 | Journal of Biomaterial Applications | 2.365 | 10.1177/0885328213519835 |
| Penghui Feng | | M 1.0 | China | Jichun Tan | M | | China | 2018 | Stem Cell Reviews and Reports | 4.924 | 10.1007/s12015-018-9867-0 |
| Peng Sun | | M 0.84 | China | Hao Wang | | M 0.89 | China | 2016 | Journal of Translational Medicine | 4.155 | 10.1186/s12967-016-0782-3 |
| Yiming Zhao | | M 0.82 | China | Hao Wang | | M 0.89 | China | 2018 | Stem Cells International | 3.937 | 10.1155/2018/3475137 |
| Man-Jing Zhang | | F 1.0 | China | Kai-Hua Lu | | M 0.86 | China | 2009 | Medical Hypotheses | 1.700 | 10.1016/j.mehy.2008.10.021 |
| Xiaoxi Xu | | M 0.56 | China | Hao Wang | | M 0.89 | China | 2018 | Stem Cell Research & Therapy | 4.777 | 10.1186/s13287-018-0874-5 |
| Michael P. Murphy | M | | USA | Niel H. Riordan | M | | USA | 2008 | Journal of Translational Medicine | 3.361 | 10.1186/1479-5876-6-45 |
| Xiuhui Chen | | F 0.86 | China | Meimei Liu | | F 0.96 | China | 2016 | International Journal of Molecular Medicine | 2.579 | 10.3892/ijmm.2016.2593 |
| Mohammad-Reza Shokri | M | | Iran | Amir-Hassan Zarnar | M | | Iran | 2019 | Scientific Reports | 4.149 | 10.1038/s41598-019-46316-3 |
| Xinxin Zhu | F | | China | Lijun Ding | | F 0.51 | China | 2019 | Current Stem Cell Research & Therapy | 2.614 | 10.2174/1574888X14666181205120110 |
| Rafael Moreno | M | | Spain | Ramon Alemany | M | | Spain | 2018 | Molecular Cancer Therapeutics | 5.151 | 10.1158/1535-7163.MCT-18-0431 |
| Lijun Chen | M Foto | F 0.51 | China | Charlie Xiang | M | | China | 2019 | Stem Cell Research & Therapy | 5.081 | 10.1186/s13287-018-1105-9 |
| Yun-xia Zhao | F | | China | Shu Lin | | F 0.57 | Australia | 2019 | Stem Cell International | 3.873 | 10.1155/2019/9071720 |
| Lijun Chen | M Foto | F 0.51 | China | Charlie Xiang | M | | China | 2019 | Stem Cell Research & Therapy | 5.081 | 10.1186/s13287-019-1503-7 |
| Aneta Sciezynska | F | | Poland | Jacek Malejczyk | M | | Poland | 2019 | Journal of Clinical Medicine | 5.583 | 10.3390/jcm8091468 |
| Siwen Zhang | M | | China | Jichun Tan | M | | China | 2019 | Stem Cell Research & Therapy | 5.081 | 10.1186/s13287-019-1155-7 |
| Qian-Yu Liu | | F 0.7 | China | Quan-Wen Liu | | M 1.0 | China | 2019 | Stem Cell International | 3.873 | 10.1155/2019/9280298 |
| Yi-Chen Wu | (foto | M 0.6 | China | Charlie Xiang | M | | China | 2019 | Stem Cell Research & Therapy | 5.081 | 10.1186/s13287-019-1243-8 |
| Marjan D. Manshadi | F | | Iran | Mehdi Abbasi | M | | Iran | 2018 | Microscopy Research and Technique | 1.355 | 10.1002/jemt.23120 |
| Yanli Liu | | F 0.87 | China | Nan Ma | F | | Germany | 2019 | Cellular and Molecular Life Sciences | 6.484 | 10.1007/s00018-019-03019-2 |
| Parastoo Noory | F | | Iran | Mehdi Abbasi | M | | Iran | 2019 | Cellular Reprogramming | 1.682 | 10.1089/cell.2019.0020 |
| Pan-Pan Cen | | M 0.62 | China | Lan-Juan Li | | F 1.0 | China | 2019 | World Journal of Gastroenterology | 3.602 | 10.3748/wjg.v25.i41.6190 |
| Elahe Mahdipour | F | | Iran | Nona Sabeti | | F 0.95 | Iran | 2019 | Journal of Cellular Physiology | 5.546 | 10.1002/jcp.28631 |
| Yang Guo | | M 0.64 | China | Charlie Xiang | M | | China | 2019 | Stem Cell and Development | 3.145 | 10.1089/scd.2018.0222 |
| Paula Barlabé | F | | Spain | Rafael Moreno | M | | Spain | 2019 | Cancer Gene Therapy | 4.472 | 10.1038/s41417-019-0110-1 |
| Hanna Manley | F | | UK | Philip Breedon | M | | UK | 2019 | Journa of Women's Health | 2.332 | 10.1089/jwh.2019.7745 |
| Song Chen | | M 0.69 | China | Jian Chen | | M 0.9 | China | 2019 | In Vitro Cellular & Develoopmental Biology - Anim | 1.587 | 10.1007/s11626-018-0311-1 |
| Han Li | | M 0.73 | Malaysia | Juntang Lin | | M 1.0 | China | 2019 | Frontiers in Molecular Neuroscience | 4.161 | 10.3389/fnmol.2019.00080 |
| Razieh Dalirfardouei | F | | Iran | Elahe Mahdipour | F | | Iran | 2019 | Journal of Tissue Enguneering and Regenerative | 3.241 | 10.1002/term.2799 |
| Pavel Deryabin | M | | Russia | Aleksandra Borodkir | F | | Russia | 2019 | Cell Cycle | 3.699 | 10.1080/15384101.2019.1593650 |
| Mina Fathi-Kazerooni | F | | Iran | Gholamreza Tavoos | M | | Iran | 2019 | Avicenna Journal of Medical Biotechnology | 1.22 | PMID: 31908739 |
| Andrea Patrizia Salzman | F | | Switzerland | Cordula Haas | F | | Switzerland | 2019 | Forensic Science International: Genetics | 5.786 | 10.1016/j.fsigen.2019.102149 |
| Akos Dobay | M | | Switzerland | Natasha Arora | F | | Switzerland | 2019 | Forensic Science International: Genetics | 5.786 | 10.1016/j.fsigen.2019.02.010 |
| Ganggang Shi | | M 0.94 | China | Hao Wang | | M 0.89 | China | 2019 | American Journal of Translational Research | 3.375 | PMID: 31497192 |
| Manijeh Khanmohammad | F | | Iran | Somaieh Kazemneja | F | | Iran | 2019 | Tissue Engineering and Regenerative Medicine | 3.241 | 10.1007/s13770-019-00189-9 |
| Zhongrui Yan | | M 1.0 | China | Xue Du | F | | China | 2019 | Stem Cell Research & Therapy | 5.081 | 10.1186/s13287-018-1101-0 |
| Penghui Feng | | M 1.0 | China | Jichun Tan | | M 0.67 | China | 2019 | Stem Cell Reviews and Reports | 5.353 | 10.1007/s12015-018-9867-0 |
| Shouyu Wang | | F 0.5 | China | Yiping Hou (M - news) | | M 0.53 | China | 2019 | Forensic Science International: Genetics | 4.884 | 10.1016/j.fsigen.2019.01.002 |
| Marine Legrand | F | | France | Anais Tondeur | F | | France | 2019 | Journal International de Bioethique et D'ethique d | dossie | 10.3917/jibes.304.0019 |
| Mina Fathi-Kazerooni | F | | Iran | Gholamreza Tavoosidana | | M 0.99 | Iran | 2019 | Biologicals | 1.795 | 10.1016/j.biologicals.2019.02.002 |
| Sook Young Yoon | | F 0.92 | Korea | autor único | | F 0.92 | Korea | 2019 | Clinical and Experimental Reproductive Medicine | 1.688 | 10.5653/cerm.2019.46.1.1 |
| Alireza Ghanavatinejad | M | | Iran | Amir-Hassan Zarnar | M | | Iran | 2020 | Journal of Reproductive Immunology | 3.535 | 10.1016/j.jri.2020.103252 |
| Ebrahim Mirzadegan | M | | Iran | Somaieh Kazemneja | F | | Iran | 2020 | International Immunopharmacology | 3.943 | 10.1016/j.intimp.2020.106595 |
| Celia Díez López | F | | Netherlands | Manfred Kayser | M | | Netherlands | 2020 | Forensic Science International: Genetics | 4.117 | 10.1016/j.fsigen.2020.102280 |
| Paula Barlabé | F | | Spain | Rafael Moreno | M | | Spain | 2020 | Cancer Gene Therapy | 4.883 | 10.1038/s41417-019-0110-1 |
| Xin Chen | | F 0.52 | China | Charlie Xiang | M | | China | 2020 | Stem Cell Research & Therapy | 5.985 | 10.1186/s13287-020-01926-x |
| Rocio Martínez-Aguilar | | F 0.99 | Spain | Ana C. Abadía-Molir | F | | Spain | 2020 | Scientific Reports | 4.13 | 10.1038/s41598-020-78423-x |
| Limei Chen | | F 1.0 | China | Long Sui | | M 0.93 | China | 2020 | American Journal of Translational Research | 3.827 | PMID:33042399 |
| Shaghayegh Arasteh | F | | Iran | Somaieh Kazemneja | F | | Iran | 2020 | Stem Cell Nanotechnology (Book) | book | 10.1007/7651_2018_193 |
| Zidong Liu | | M 1.0 | China | Gengqian Zhang | | M (Foto or | China | 2020 | Electrophoresis Journal | 3.062 | 10.1002/elps.202000053 |

| | | | | | | | | | | | |
|---|---|---|---|---|---|---|---|---|---|---|---|
| Mahmood Bozorgmehr | M | | Iran | Caroline E. Gargett | F | | Australia | 2020 | Frontiers in Cell and Developmental Biology | 5.868 | 10.3389/fcell.2020.00497 |
| Rosana de Almeida Sant | F | | Brazil | Regina Coeli dos Sa | F | | Brazil | 2020 | International Journal of Molecular Sciences | 5.542 | 10.3390/ijms21249563 |
| Sedighe Esmaeilzadeh | F | | Iran | Mohammad Ghasen | M | | Iran | 2020 | The Journal of Obstetrics and Gynaecology Resea | 1.524 | 10.1111/jog.14340 |
| Shanti Gurung | F | | Australia | Caroline E. Gargett | F | | Australia | 2020 | Journal of Personalized Medicine | 1.462 | 10.3390/jpm10040261 |
| Alicia Sanzhez-Mata | F | | Spain | Elena Gonzalez-Mu | F | | Spain | 2020 | iScience | 5.080 | 10.1016/j.xpro.2020.100183 |
| Jiajia Chen | | F 0.78 | China | Lanjuan Li | | F 1.0 | China | 2020 | Engineering | 6.495 | 10.1016/j.eng.2020.02.006 |
| Lidia Lopez-Caraballo | F | | Spain | Elena Gonzalez-M | F | | Spain | 2020 | iScience | 5.080 | 10.1016/j.isci.2020.101376 |
| Wang Jin | | M 0.67 | China | Hao Wang | | M 0.89 | China | 2020 | Stem Cells International | 4.72 | 10.1155/2020/4820543 |
| Wenchun Qu | M | | USA | Fred P. Sanfilippo | M | | USA | 2020 | Stem Cells Translational Medicine | 5.607 | 10.1002/sctm.20-0146 |
| Marianna Ferreira Gonça | F | | Brazil | Regina Coeli dos Sa | F | | Brazil | 2020 | Tissue Engineering | 3.312 | 10.1089/ten.tea.2020.0034 |
| Qi-Yuan Chang | | M 0.83 | China | Ji-Chun Tan | | M 0.67-0.7 | China | 2020 | World Journal of Stem Cells | 5.326 | 10.4252/wjsc.v12.i5.368 |
| Simin Zafardoust | | F 0.88 | Iran | Afsaneh Mohammadzadeh | | F 0.98 | Iran | 2020 | Stem Cell Reviews and Reports | 4.813 | 10.1007/s12015-020-09969-6 |
| Hyun Sok Chung | | M 0.63 | South Korea | Autor único | | M 0.63 | | 2019 | Korean Journal of Medical History | 0.12 | 10.13081/kjmh.2019.28.239 |
